# Roles of meteorological conditions in COVID-19 transmission on a worldwide scale

**DOI:** 10.1101/2020.03.16.20037168

**Authors:** Biqing Chen, Hao Liang, Xiaomin Yuan, Yingying Hu, Miao Xu, Yating Zhao, Binfen Zhang, Fang Tian, Xuejun Zhu

## Abstract

The novel coronavirus (SARS-CoV-2/ 2019-nCoV) identified in December 2019 has caused great damage to public health and economy worldwide. Previous research has suggested an involvement of meteorological conditions in the spread of droplet-mediated viral diseases, such as influenza. However, as for the recent novel coronavirus, few studies have discussed systematically about the role of daily weather in the epidemic transmission of the virus. Here, we examine the relationships of meteorological variables with the severity of the outbreak on a worldwide scale. The confirmed case counts, which indicates the severity of COVID-19 spread, and four meteorological variables, i.e., air temperature, relative humidity, wind speed, and visibility, were collected daily between January 20 and March 11 (52 days) for 430 cities and districts all over China, 21 cities/ provinces in Italy, 21 cities/ provinces in Japan, and 51 other countries around the world. Four different time delays of weather (on the day, 3 days ago, 7 days ago, and 14 days ago) as to the epidemic situation were taken formodeling and we finally chose the weather two weeks ago to model against the daily epidemic situation as its correlated with the outbreak best. Taken Chinese cities as a discovery dataset, it was suggested that temperature, wind speed, and relative humidity combined together could best predict the epidemic situation. The meteorological model could well predict the outbreak around the world with a high correlation (*r*^*2*^ > 0.6) with the real data. Using this model, we further predicted the possible epidemic situation in the future days for several high-latitude cities with potential outbreak. This model could provide more information for government’s future decisions on COVID-19 outbreak control.

## INTRODUCTION

In the first season of 2020, an outbreak of atypical pneumonia (COVID-19) caused by a novel coronavirus (SARS-CoV-2 or 2019-nCoV)^1^ has spread all over the world and had a great impact on public health and worldwide economy. This new virus has some relations to SARS-CoV but it is more aggressive than SARS, MERS, or the seasonal influenza^2^. It has exhibited relatively high human-to-human transmissibility compared to other coronavirus infections^3^.

Although the outbreak of COVID-19 in China was controlled by rigorous blockade and isolation, the outbreak around the world is still an important public health problem and leads to worldwide economic crisis. For government decision making, it is helpful to know the prediction of the future trend of COVID-19 outbreak ahead.

To predict epidemic trend, meteorological conditions are suggested to be an important factor as well as population mobility and human-to-human contact. These meteorological factors such as humidity, visibility, and wind speed can affect droplet stability in the environment, or affect survival of viruses as air temperature does, and thus impact epidemic transmission. Air temperature and absolute humidity have been indicated to significantly influence the transmission of COVID-19. However, there are only a few studies published on preprint hubs discussed the involvement of meteorological conditions in the spread of COVID-19 up to now, and they all focused on data from several cities without model validation or prediction.

Herein, this study intends to investigate the relationship between meteorological factors and epidemic transmission rate at a systematical level on the world scale. Four meteorological variables, i.e., air temperature, relative humidity, wind speed, and visibility, were collected as well as the confirmed case counts for each day between January 20 and March 11 for 430 cities and districts all over China. Those cities with over 50 confirmed cases monthly were taken as a discovery dataset to exclude the confounding effect due to purely imported cases. Four time points delay of the weather conditions from the day of epidemic situation evaluation were considered and compared to find the most possible time delay that best reveals the relationship between weather and COVID-19 outbreak. A multivariate polynomial regression model with factors of wind speed, relative humidity, and average air temperature was established, and then validated in 21 cities/ provinces in Italy, 21 cities/ provinces in Japan, and 51 other countries around the world. Finally, we use this model, in combination with weather forecast, to predict the probable outbreak in several high-latitude big cities around the world.

## MATERIAL AND METHODS

### Epidemiological data

Epidemiological data were collected from various sources, including the World Health Organization (WHO)^4^, China Center for Disease Control and Prevention CDC, European Centre for Disease Control and Prevention (especially for data in Italy), Japan Center for Disease Control and Prevention, and DXY-COVID-19-Data, a Chinese website that aggregates national and local CCDC situation reports in near real-time^5^. The daily new confirmed case counts were collected from January 20, 2020 (i.e., WHO published the first situation report) to March 11, 2020. For China, incidence data were collected for every city or district, leading to 430 sites covering the whole country. Considering the potential confounding effect, only cities with no less than 50 cumulative confirmed cases in one month and without official reports of imported cases in majority were taken into a discovery dataset. A second set of cities with 10∼49 cumulative confirmed casesin one month was taken as an replication dataset (so called replication_China). For Italy and Japan, incidence data were collected for each province/ city/ district, in total, 21 sites for Italy (replication_Italy) and 21 for Japan (replication_Japan). For other countries, only incidence data at the country level were obtained. We scrutinized WHO’s situation reports to rule out these countries with only imported cases, and only collected the confirmed cases with possible or confirmed local transmission (i.e., without recent travel history to China). The top 11 countries with high incidence of COVID-19 local transmission except China were selected for a replication dataset representing the world’s situation (so called replication_world).

### Weather data

We obtained hourly values of meteorological observations from the Integrated Surface Database of USA National Centers for Environmental Information (NCEI, https://www.ncdc.noaa.gov/isd)^6^. Temperature and dew point displayed in Fahrenheit were transformed into Celsius forms, and relative humidity (RH) was calculated from temperature and dew point using the following formula for each time point:

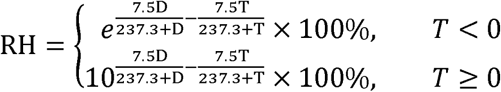

where RH is the relative humidity, D is the dew point in degrees Celsius, T is the temperature in degrees Celsius, and *e* is the base of the natural log.

Daily data were calculated by averaging the hourly data for each variable in each day. For each city with epidemiological data, the meteorological station in that city or that was closest to the latitude and longitude coordinates of the city center was chosen. For a city with more than one meteorological stations, the one nearest to the city center was chosen. For a province with epidemiological data, the meteorological station in the capital city of that province was chosen. For a country with only nation-wide epidemiological data, weather data were averaged across all the meteorological stations in the cities where outbreak was officially reported. For Japan, the surveillance table of epidemiology only provides weekly data, thus daily weather of each week was averaged to obtain weekly meteorological data.

### Statistical modeling

The number of confirmed new cases on each day was taken as a dependent variable.Four meteorological variables, namely, air temperature, wind speed, visibility, and relative humidity, were taken as independent variables. Considering that there is a latency stage from the day one get infected to the day being confirmed, a time delay of the day COVID-19 was confirmed from the day weather data were collected needs to be taken into consideration. As it is reported that the latency period for COVID-19 is 3∼7 days on average and 14 days at most, four time points delay of virus infection were taken into consideration, that is, weather data were collected on the day, three days before, seven days before, 14 days before collecting the epidemiological data. At first, each meteorological variablewas plotted against the confirmed new case counts for the Wuhan dataset, with four time delays display on one plot. Only one city Wuhan was chosen for illustrating the time delay effectbecause it is the original city where SARS-CoV-2 was first uncovered, there could not be any imported cases for Wuhan, which might obscure the correlation between weather and virus transmission. A Loess regression interpolation approach was adopted to visually identify the relationship between meteorological variables and confirmed new case counts. After choosing the appropriate time delay, data from the discovery dataset were fitted into generalized linear model or non-linear model (basically polynomial and inverse models) according to the indentified relationship by Loess regression. Each of the four meteorological variables was fitted into models solely, and then two or three variables were combinedtogether to fit complex models. All these models were compared with the full model, to find a best fitted model with not many factors. Basic statistics and modeling was conducted in R 3.5.1^7^ (mainly “glm”, “nls”, “ggplot” packages).

### Model validation and application

The best fitted model was validated in the replication datasets (replication_world, replication_Italy, and replication_Japan) by correlating the real epidemiological data with the predicted values from the model.

## RESULTS

There were in total 39,888 confirmed cases in Wuhan, and 14,511 confirmed cases in 59 Chinese cities /districts with monthly confirmed cases no less than 50, therefore, the discovery dataset was consisted of 1,133 records of data with 54,399 confirmed cases in 60 cities in China. The confirmed new cases in Wuhan on February 13, 2020, reached 13,436, which was oddly high as the daily confirmed new cases were no larger than 3,000 on all the other dates in Wuhan or in all the other cities. We suppose that it might be due to supplement of enough virus test kits on that day. In order to reduce the potential contamination of modeling by this outlier, we substituted the counts on that day by four, that was 13,436/4=3,359, which was still the largest number but not deviated from the dataset too much. Except this outlier, the daily confirmed new cases in the discovery dataset ranged from 1 to 2997, the average temperature ranged −23.54°C ∼ 22.85°C, the wind speed ranged 1.33 ∼ 26 miles per hour, visibility ranged 0.425 ∼ 110 statute miles to nearest tenth, and relative humidity ranged 31.4% ∼ 100%. The temperature, wind speed, and relative humidity ranges in the other replication datasets were similar to the discovery dataset, while the maximum visibility in the replication datasets was lower than that in the discovery dataset (Table 1).

**Table 1.**
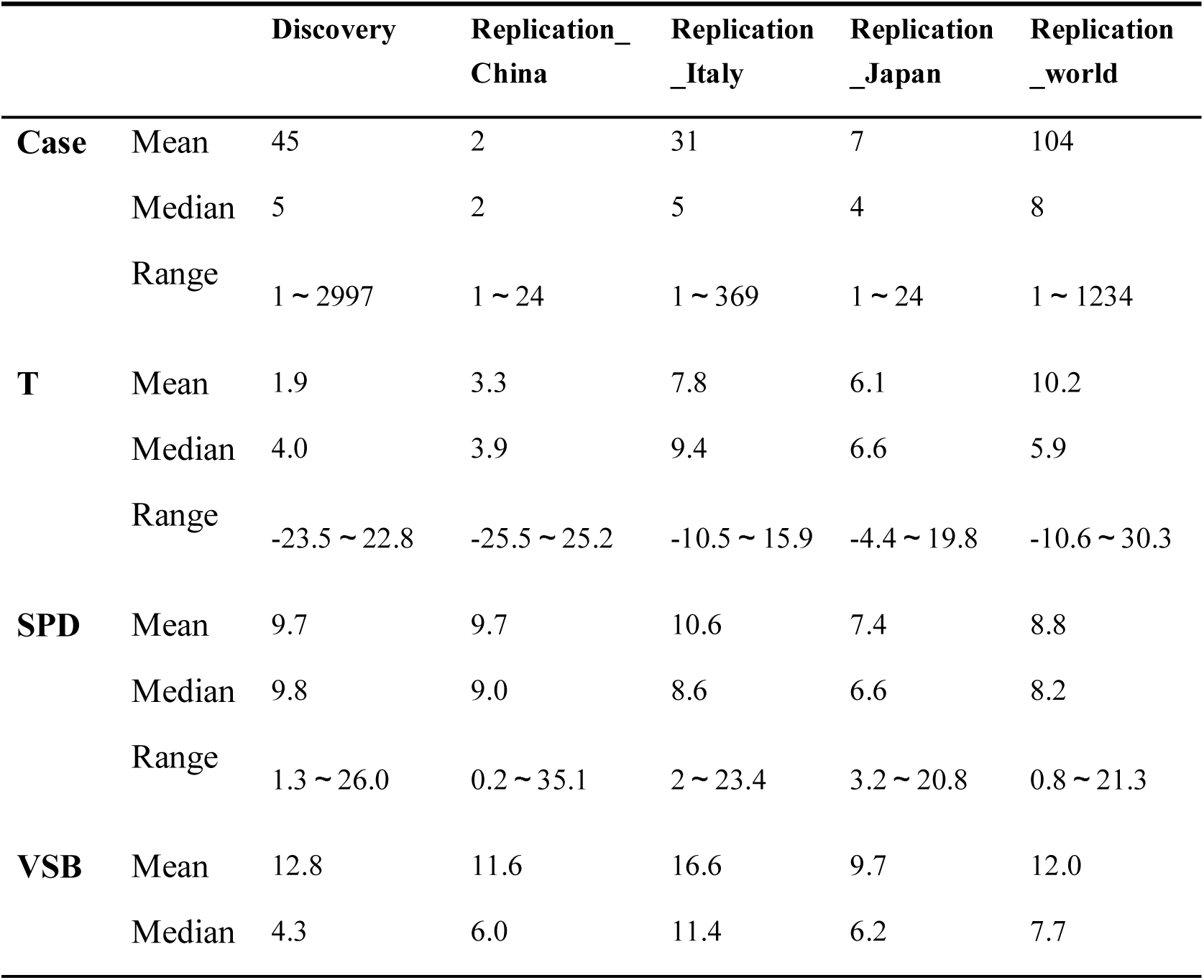
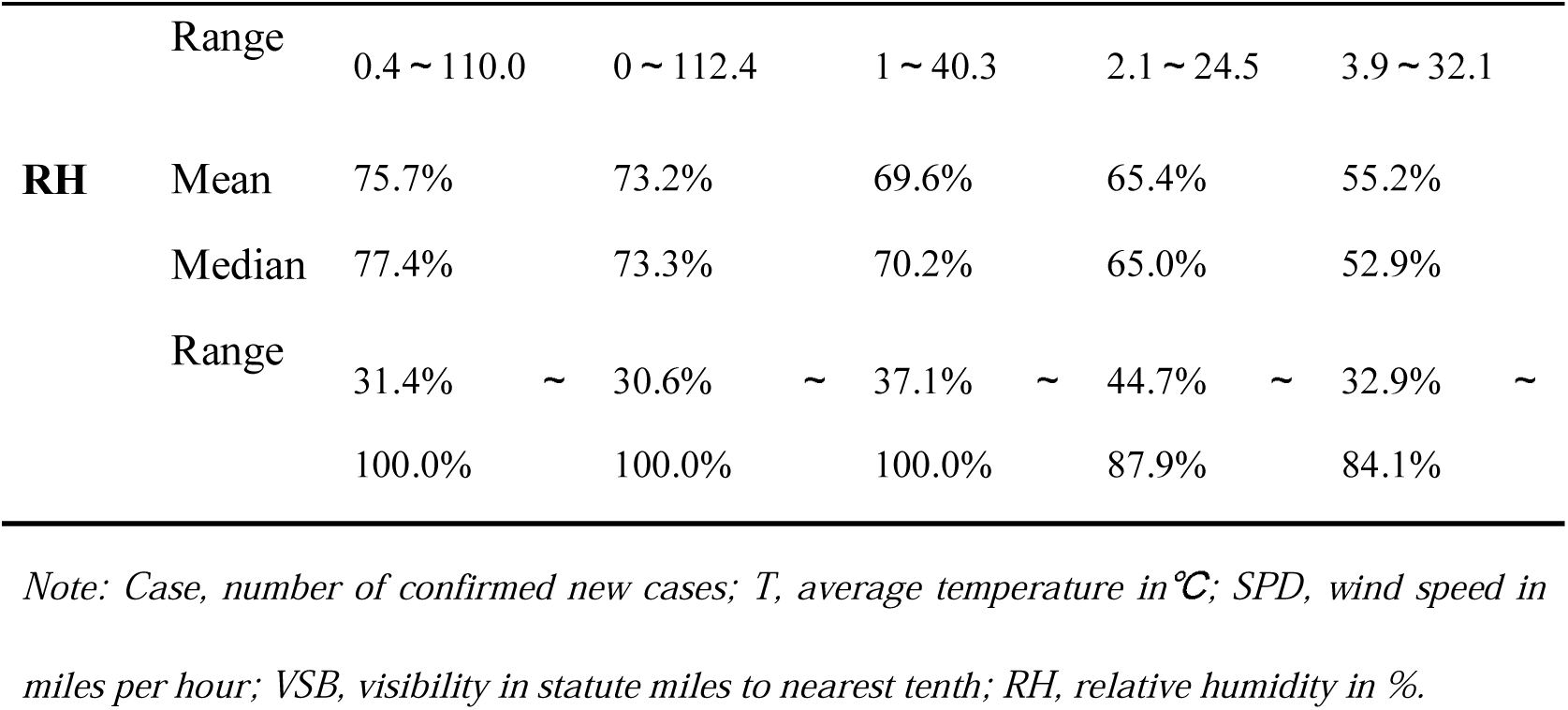
Basic summary statistics of the epidemiological and meteorological data.

Regression interpolation showed that the weather two weeks ago was correlated with the confirmed new case counts in a most reasonable manner for temperature, relative humidity, and visibility. The effects of all these four factors on confirmed new cases 14 days later all exhibited a parabolic or bell-shaped trend (Figure 1). Thus, in the following analyses, epidemiological data were correlated with the weather data 14 days ago.

**Figure 1.**
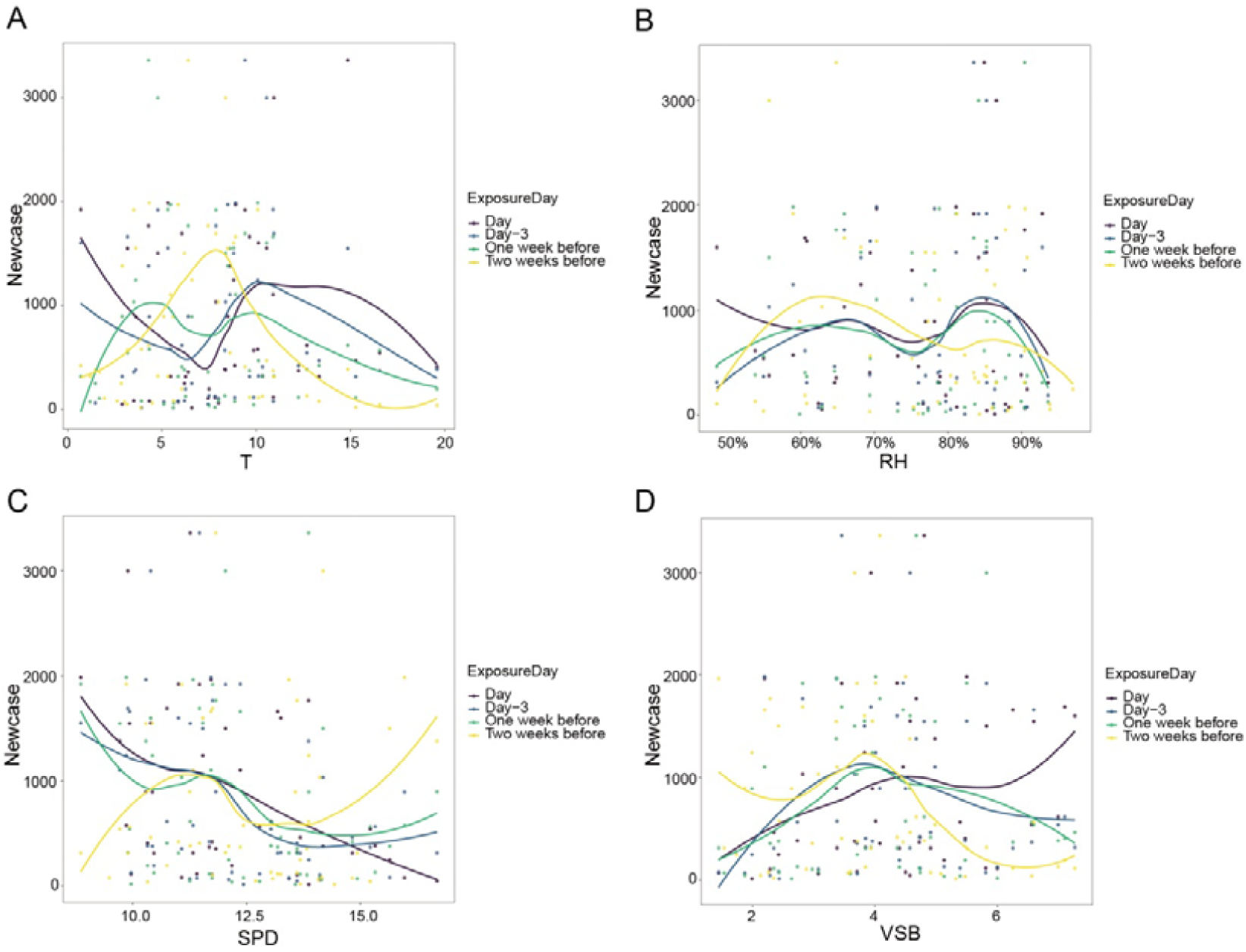
Loess regression interpolation of confirmed new case counts to the four meteorological variables, (A) average temperature (T) in °C, (B) relative humidity (RH) in %, (C) wind speed (SPD) in miles per hour, (D) visibility (VSB) in statute miles to nearest tenth, for Wuhan city. Four time delay of the confirmation day (when epidemiological data were correlated) from the exposure day (when weather data was correlated) are displayed together in one figure, namely, exposure on the day, three days before, one week before, two weeks before.

The correlation between weather and epidemic situation showed similar patterns for the main outbreak cities in China other than Wuhan and for the other outbreak countries (Figure 2).

**Figure 2.**
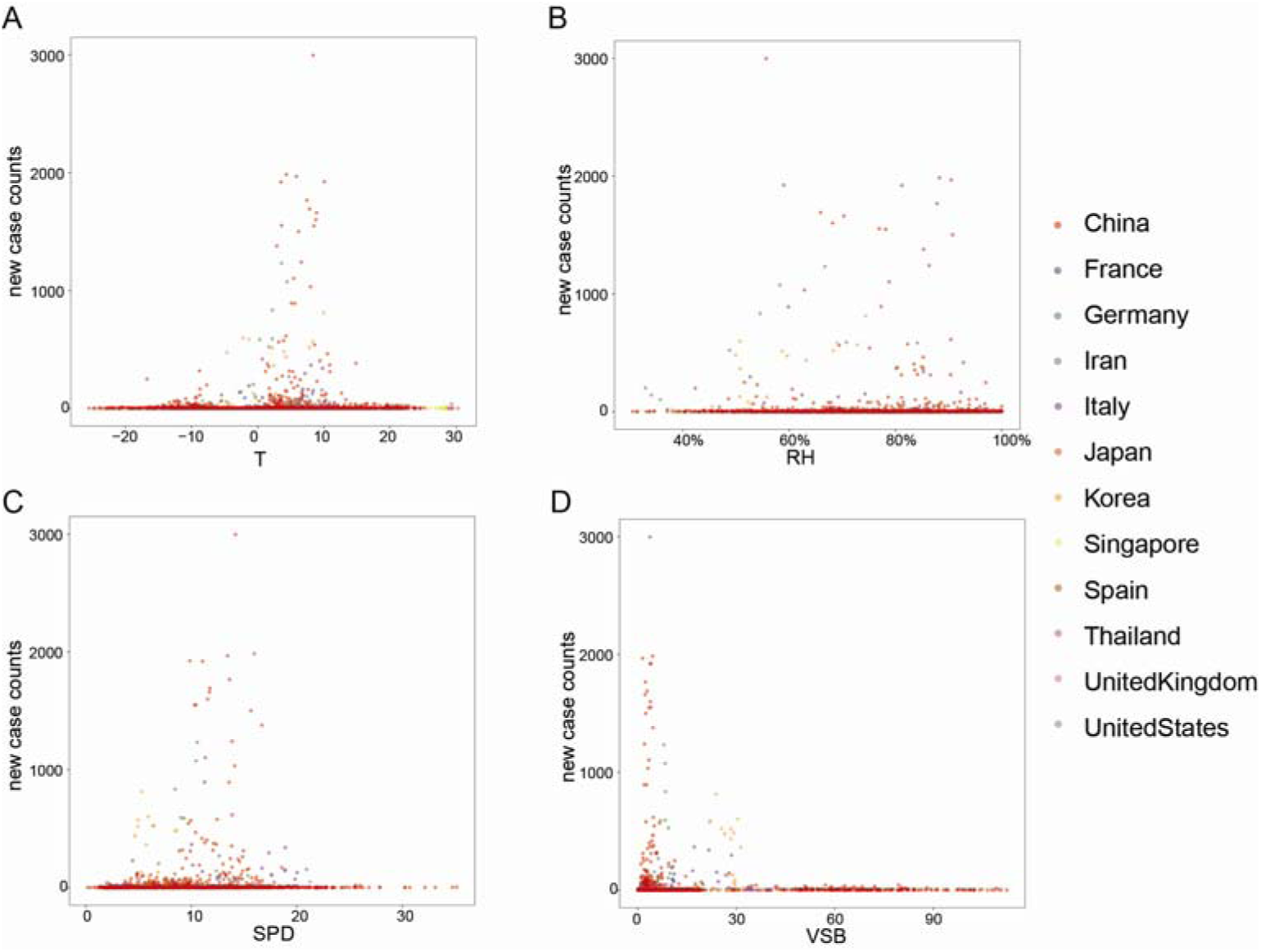
Scatterplots of confirmed new case counts to the four meteorological variables, (A) average temperature (T) in °C, (B) relative humidity (RH) in %, (C) wind speed (SPD) in miles per hour, (D) visibility (VSB) in statute miles to nearest tenth, for all the studied sites in the top 12 outbreak countries.

Loess regression interpolation for each dataset showed that the relationship between weather and epidemic situation in each replication dataset was similar to that in the discovery dataset, all the four meteorological variables exhibited a bell-shaped trend, in detail, the relationship looks quadratic for temperature, wind speed, and visibility, and cubic for relative humidity (Figure 3).

**Figure 3.**
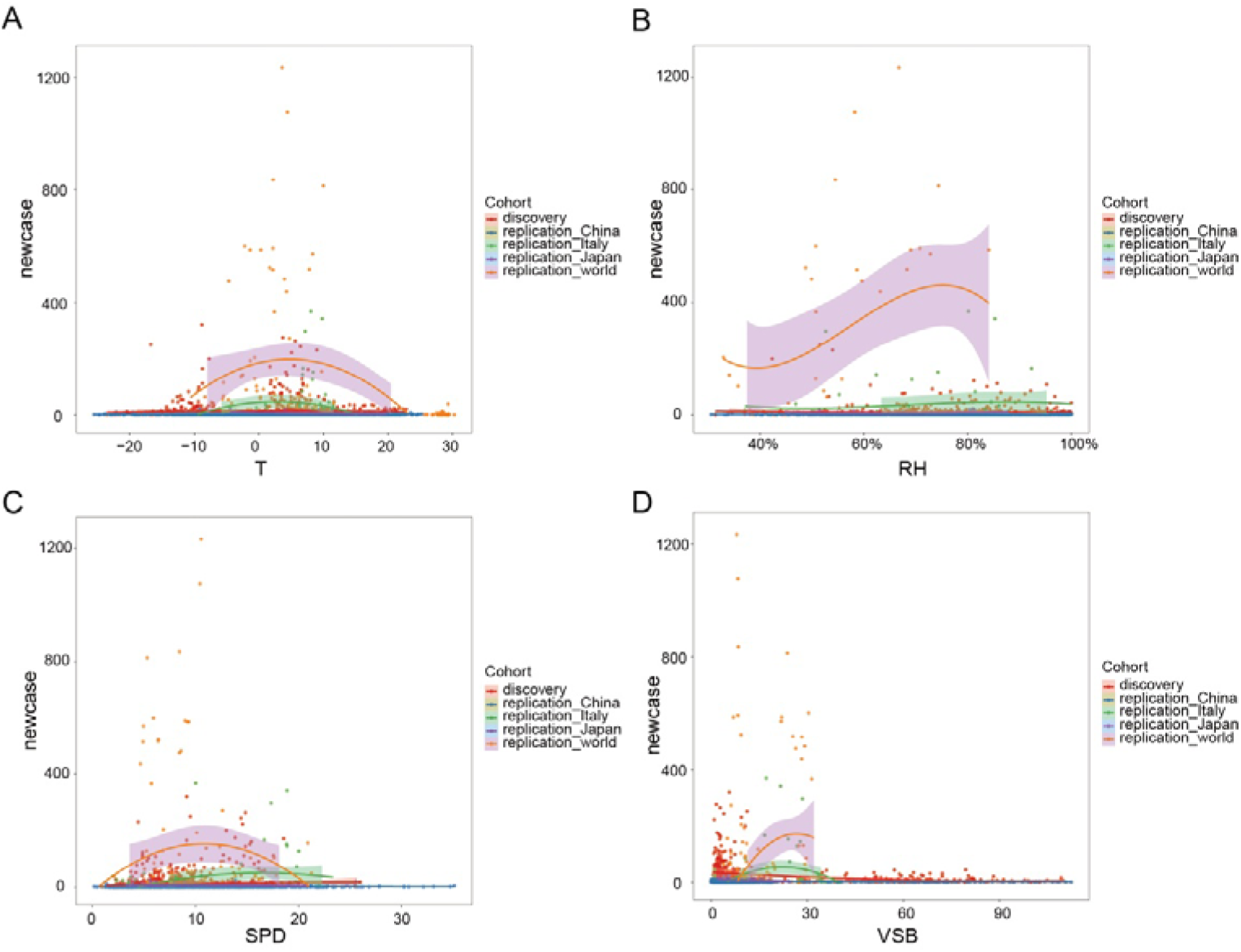
Scatterplots of confirmed new case counts to the four meteorological variables, (A) average temperature (T) in °C, (B) relative humidity (RH) in %, (C) wind speed (SPD) in miles per hour, (D) visibility (VSB) in statute miles to nearest tenth, for all the studied datasets. Loess regression interpolation curves with 95% confidence intervals in shadow were illustrated for each dataset.

To elucidate the contribution of each meteorological factor to the case counts, we first performed single-factor non-linear regression modeling for each meteorological variable in the Wuhan dataset as well as in the discovery dataset. Temperature and wind speed were fitted into quadratic models; relative humidity was fitted into a cubic model; visibility was fitted into two models, an inverse model when modeling in the discovery dataset and a quadratic model when modeling in the Wuhan dataset because distribution of visibility in the two datasets was different. We used these fitted models to calculate a predicted value for case counts for each studied site, and then compared this predicted value with the real observed case counts by calculating a Pearson’s correlation coefficient between them. Model fitting results showed that using the Wuhan dataset for single-factor modeling produced better model fitness. There was 0.40, 0.24, and 0.35 correlation between the observed data for Wuhan and values predicted by average air temperature, relative humidity, and visibility, separately, while wind speed alone could not explain much of the variance in confirmed case counts (Figure 4). According to the equation, SARS-CoV-2 transmission reaches a peak when the air temperature is 8.07 °C, or when the wind speed is 16.1 mile/hr, or when the visibility is 2.99 statute miles to nearest tenth, or when the relative humidity is 64.6%.

**Figure 4.**
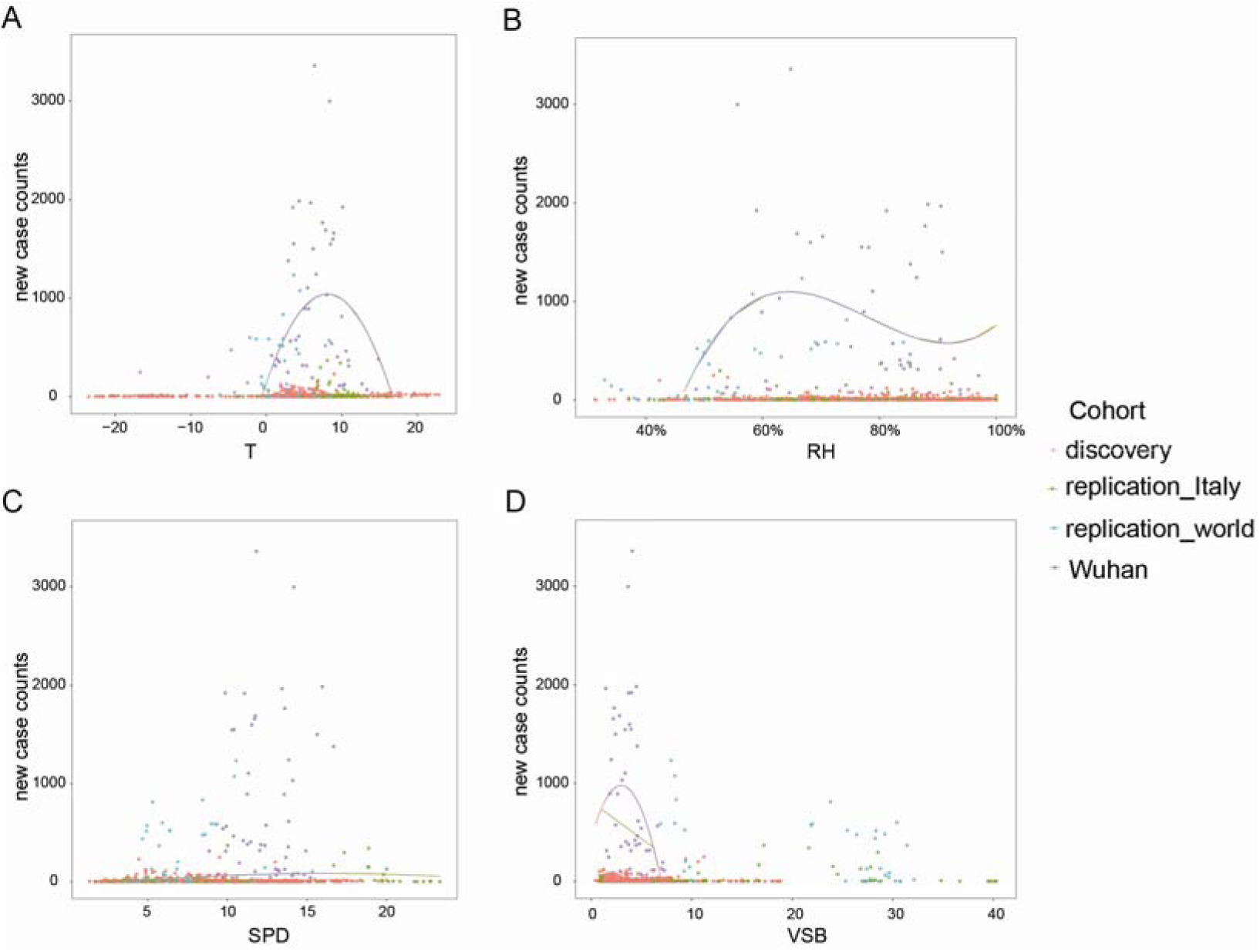
Regression curves on each dataset, showing the best fitted single factor model for each meteorological variable, (A) average temperature (T) in °C, (B) relative humidity (RH) in %, (C) wind speed (SPD) in miles per hour, (D) visibility (VSB) in statute miles to nearest tenth. The fitted models are (A) case counts ∼ -13.826T^2^ + 223.111T + 140.958, (B) case counts ∼ 52964RH^3^ -124085RH^2^ + 94004RH-22124, (C) case counts ∼ -0.5458SPD^2^17.6353SPD -58.4365, (D) case counts ∼ -61.29VSB^2^ + 366.03VSB +432.43.

As the reality is that a single weather factor alone could not affect the virus transmission too much, we further combined different meteorological variables to fit a more complex model, in order to take the systematic influence by different types of weather data into consideration. To fit the model with more data and thus more accuracy, we used the discovery dataset for modeling. In the model, temperature and wind speed were regarded as quadric-related, relative humidity was regarded as cubic-related, and visibility was regarded as inverse-related. The full model fitted was as follows:

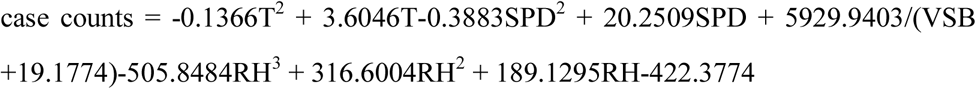

where, T is in □, SPD is in miles per hour, VSB is in statute miles, RH is in decimal. Using this full model for prediction in the replication datasets, we got a quite good prediction result for the national data all over the world (replication_world), with a case counts prediction significantly correlated with the real data (Pearson’s correlation coefficient *r*^*2*^ = 0.487, *p* = 0.003; Figure 5A). When further reducing variables in the model to obtain a most parsimony and best fitted prediction model, we got better results. When visibility was removed from the model, the predicted values of the fitted model were more significantly correlated with the observed epidemiological data (*r*^*2*^ = 0.624, *p* = 6.113e-05 for replication_world; *r*^*2*^ = 0.287, *p* = 0.034 for replication_Italy; see Figure 5B & Figure 6). This model, written as follows, was also best fitted compared to the full model and other 3-factor and 2-factor models, with the smallest Akaike information criterion.

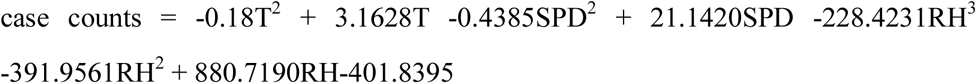

where, T is in °C, SPD is in miles per hour, VSB is in statute miles, RH is in decimal.

**Figure 5.**
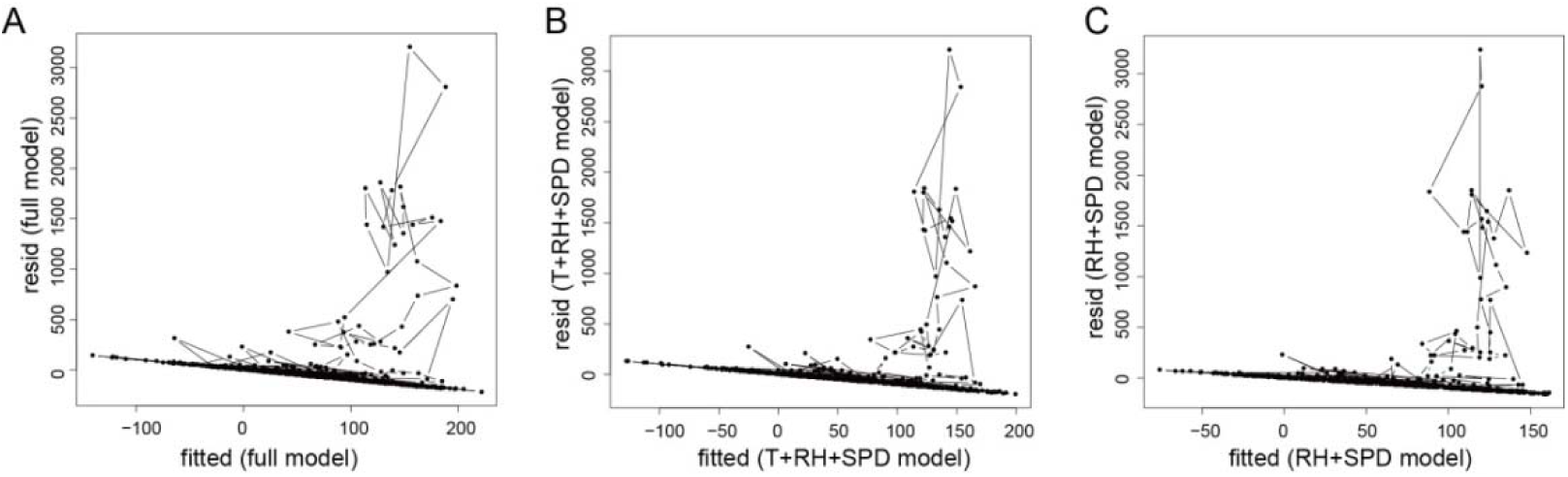
Residues versus fitted values plots for (A) the full model; (B) the 3-factor model with temperature (T), relative humidity (RH), and wind speed (SPD); (C) the 2-factor model with relative humidity and wind speed.

**Figure 6.**
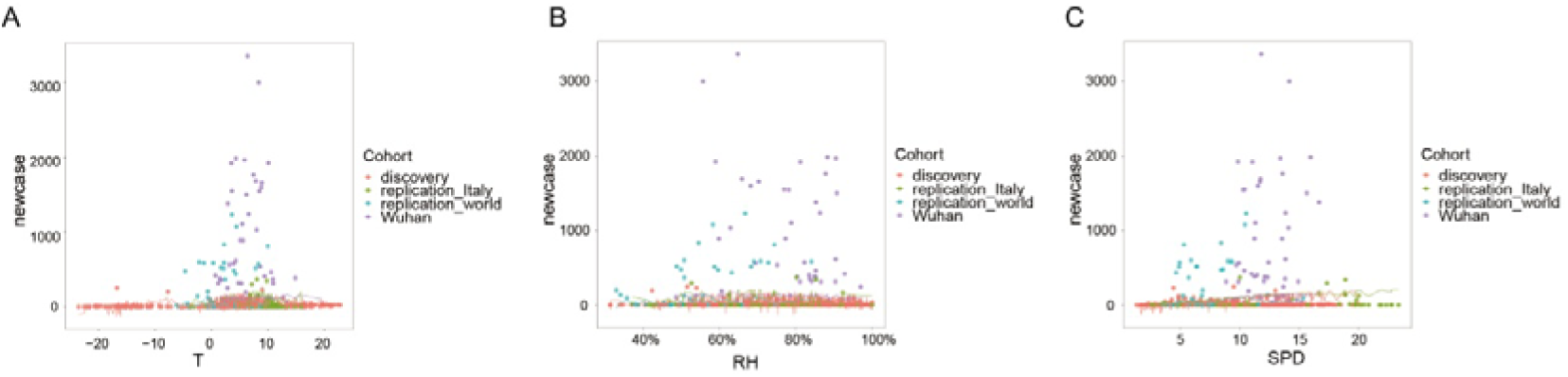
Fitted curve of the best fitted multivariate model, projected to each meteorological variable. Lines illustrate the change in predicted values by the best-fitted model as (A) temperature, (B) relative humidity, and (C) wind speed changes. Dots represent data set in each studied site.

When there were only wind speed and relative humidity in the model, we got a prediction that was most significantly correlated with the real data (*r*^*2*^ = 0.637, *p* = 3.884e-05 for replication_world; *r*^*2*^ = 0.310, *p* = 0.021 for replication_Italy; see Figure 5C & Figure 7), but the model fitness was not the best.

**Figure 7.**
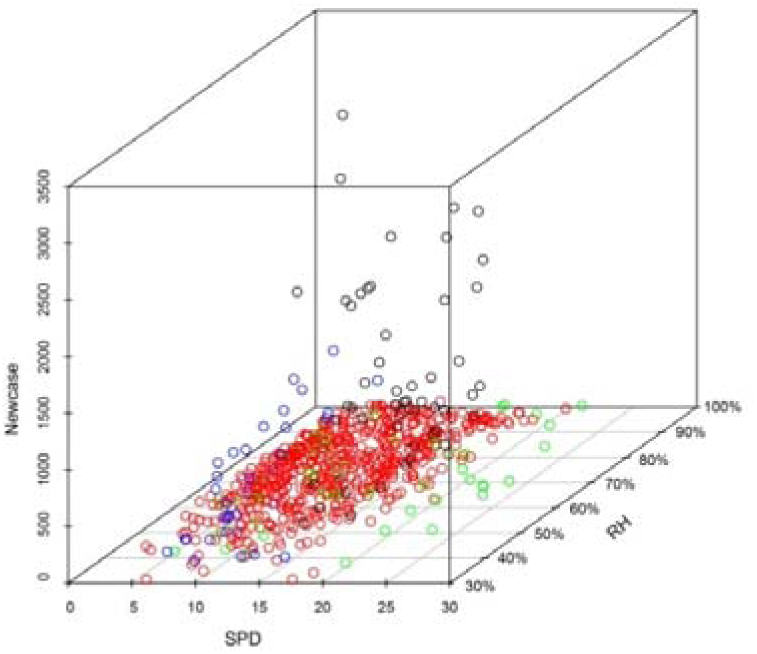
3D plot illustrating the relationship of confirmed new case counts (Newcase) with wind speed (SPD) and relative humidity (RH). Each dot represents a site. Black dots represent data of Wuhan; red dots represents data of Chinese cities in the discovery dataset other than Wuhan; green dots represents data of Italy cities; blue dots represents data of replication _world.

## DISCUSSION

Significant impact of different temperature exposure on the human-to-human transmission of COVID-19 has been reported by a few studies, absolute humidity has also been related to human-to-human transmission of COVID-19. However, there is currently no systematic and quantitative research investigating the exact impact of a set of meteorological factors on the spread rate of COVID-19. Our study suggests that changes in a single weather factor, such as temperature or humidity, could not correlate with the case counts very well. On the other hand, several meteorological factors combined together could describe the epidemic trend much better than single-factor models. Our research also finds that there exists nonlinear dose-response relationship for all the four meteorological factors, in consistency with previous studies about climate and epidemics. Predictions of COVID-19outbreak scale by the models were well correlated with the observations, suggesting an important role of weather in the transmission of SARS-CoV-2 all over the world.

The impact of weather on the spread of plague has been discussed early in human’s history. The ancient Chinese had a theory called “Five Movement and Six Weather” to study climate change and its relationship with human health and diseases. This theory is a summary of long-term observations on astronomy, astrology, calendar and meteorology. According to this theory, the year 2020 is predicted to be warm and dry, and plague often outbreaks in warm winter, so respiratory infectious diseases would be frequent in 2020. This theory and its inference are consistent with the current epidemic situation of COVID-19.

Previous studies have already implied the spread of many respiratory infectious diseases like influenza is dependent upon air temperature and relative humidity^8,9^. Recent published papers^10-14^ on the relationship of weather and COVID-19 have reported significant association of COVID-19 transmission and temperature and humidity, but their conclusions are controversial. Cai et al did not find any correlation between the growth rate of the epidemics and daily mean temperature in either Wuhan or Hunan^10^. On the contrary, our study suggests significant impact of daily mean temperature on the daily confirmed new case counts 14 days later. It is supposed that a sufficient time delay between exposure and confirmation is crucial for weather to exhibit its effect. Luo et al suggested that changes virus transmission occurred in a wide range of humidity and temperature conditions^11^ and Bu et al concluded that temperature ranging 13∼19°Cand humidity in 50% ∼ 80% are suitable for the survival and transmission of this new coronavirus^12^. However, our study suggests that there is a relatively not wide temperature and humidity ranges for SARS-CoV-2 spread, there is an optimal temperature for SARS-CoV-2 at 8.07 °C and most cities with high epidemic transmission of COVID-19 locate in the humidity range of 60% ∼ 90%, which is colder and more humid than suggestions by Bu et al. It might be due to that their conclusion was based on SARS data and their data collection was much earlier. Our optimal temperature 8.07 °C is very close to the estimation by Wang et al, which is 8.72°C13. Oliveiros et al. regressed the doubling time of COVID-19 cases by temperature and humidity, and they did not find significant association for wind speed^14^. In our study, though wind speed was not an important factor if modeled singly, it is a necessary factor in the final model and adding wind speed in the model would significantly improve model prediction performance. Another interesting thing to mention in our study is that air visibility was negatively correlated with case counts in an inverse manner, while case counts decrease rapidly when visibility is high. It suggests that caution about outbreak should be taken if visibility drops below 10 statute miles to nearest tenth. Upon now, all of these studies focused only on data from China, with very few worldwide data implemented. Our research investigated the worldwide data more thoroughly and explored a set of meteorological factors systematically. We have found the correlation with epidemic trend for not only temperature and humidity, but also wind speed and visibility. We are the first to take visibility into consideration, and it is suggested that when visibility is lower than 30 statute miles to nearest tenth, caution should be made about COVID-19 outbreak.

None of the published research has considered the influence of imported cases in modeling. In our study, when collecting epidemiological data for other countries in the world, those cases with travel history to China or indicated by WHO as “imported case only” were excluded, leaving the world data most likely local transmitted. However, it’s difficult to separate the imported cases from local transmission for Chinese cities, as there was a dramatic and complex migration due to Spring Festival. It might explain why the predicted values for Wuhan and countries other than China correlated with the observed data much better than those for Chinese cities other than Wuhan. Future research should investigate the epidemiological data more carefully and thoroughly to distinguish imported cases and local transmission.

A final prediction model is proposed in the current study, which is easy to use for estimating the future 14-day epidemic trend of COVID-19 by using weather observations in the past two weeks. However, if strict control on population movement and clustering is implemented, the real case counts might deviate from the predicted values.

## Data Availability

Data are available upon request by email.

## ACKNOWLEDGEMENT

This work is supported by the Priority Academic Program Development of Jiangsu Higher Education Institutions – the third period (NO.035062002003c), the “Yizhong” Research Grand of Jiangsu Province Hospital of Chinese Medicine (Y19066). We thank Dr. Zhisheng Huang for advices on data collecting and processing, Dr. Siyuan Tan and Dr Yonggao Chen for technical support on analyses.

## REFERENCE

1. Zhu N, Zhang D, Wang W, Li XW, Yang B, Song JD, et al. A novel coronavirus from patients withpneumonia in China, 2019. N Engl J Med, Feb 2020. 382:727–733.

2. Wang C, Hornby PW, Hayden FG, Gao GF. A novel coronavirus outbreak of global health concern. Lancet, Feb 2020. 395(10223): 470–473.

3. Chan JFW, Yuan S, Kok KH, To KKW, Chu H, Yang J, et al. A familial cluster of pneumoniaassociated with the 2019 novel coronavirus indicating person-to-person transmission: a studyof a family cluster. Lancet, Feb 2020. 395: 497–506.

4. World Health Organization. Novel coronavirus (2019-nCoV). Available from:https://www.who.int/emergencies/diseases/novel-coronavirus-2019/situation-reports [accessed on 2020-03-12]

5. https://github.com/BlankerL/DXY-COVID-19-Data [accessed on 2020-03-12]

6. ftp://ftp.ncdc.noaa.gov/pub/data/noaa/2020/ [accessed on 2020-03-16]

7. R Core Team (2017). R: A language and environment for statistical computing. R Foundation forStatistical Computing, Vienna, Austria. URL https://www.R-project.org/

8. Barreca, A.I. and J.P. Shimshack, Absolute humidity, temperature, and influenza mortality:30 years of county-level evidence from the United States. American journal of epidemiology, 2012. 176(suppl 7): p. S114–S122.

9. Lowen AC, Mubareka S, Steel J, Palese P. Influenza Virus Transmission is Dependent on RelativeHumidity and Temperature. PLoS Pathol, 2007 Oct; 3(10): e151.

10. Cai Y, Huang Sr. T, Liu Sr. X, Xu Sr. G. The Effects of “Fangcang, Huoshenshan, and Leishenshan” Makeshift Hospitals and Temperature on the Mortality of COVID-19. Preprint at Med RXIV. 2th March 2020; doi: https://doi.org/10.1101/2020.02.26.20028472

11. Luo W, Majumder MS, Liu D, Poirier C, Mandl KD, Lipsitch M, Santillana M. The role of absolutehumidity on transmission rates of the COVID-19 outbreak. Preprint at Med RXIV. 17th February 2020; doi: https://doi.org/10.1101/2020.02.12.20022467

12. Bu J, Peng DD, Xiao H, Yue Q, Han Y, Lin Y, Hu G, Chen J. Analysis of meteorological conditions andprediction of epidemic trend of 2019-nCoV infection in 2020. Preprint at Med RXIV. 18th February 2020; doi: https://doi.org/10.1101/2020.02.13.20022715

13. Wang M, Jiang A, Gong L, Luo L, Guo W, Li C, Zheng J, Li C, Yang B, Zeng J, Chen Y, Zheng K, Li H. Temperature significant change COVID-19 Transmission in 429 cities. Preprint at Med RXIV. 25th February 2020; doi: https://doi.org/10.1101/2020.02.22.20025791

14. Oliveiros B, Caramelo L, Ferreira NC, Caramelo F. Role of temperature and humidity in the modulation of the doubling time ofCOVID-19 cases. Preprint at Med RXIV. 5th March 2020; doi: https://doi.org/10.1101/2020.03.05.20031872

